# An Fc receptor and IgA functional signature identifies TB disease in children living with HIV

**DOI:** 10.64898/2026.02.08.26345833

**Authors:** Ye jin Kang, Nannan Wang, Amyn Malik, Pei Lu, Irene Njuguna, Elizabeth Maleche-Obimbo, Sylvia M. LaCourse, Jennifer Slyker, Dalton Wamalwa, Grace C John-Stewart, Chuangqi Wang, Lisa M. Cranmer, Lenette L. Lu

## Abstract

**Background:** Tuberculosis (TB) is a leading cause of morbidity and mortality among children living with HIV (CLHIV). Poor diagnostic performance is a significant contributor. Serological assays that determine levels of *Mycobacterium tuberculosis* reactive antibodies inconsistently detect TB. However, antigen-specific antibody Fc receptor engagement and effector functions are promising biomarkers of TB disease.

**Methods:** This study evaluated serum from a well-characterized cohort of Kenyan CLHIV via two orthogonal approaches: 1) longitudinally following over the course of TB treatment and 2) assessing a cross-section with and without clinical TB disease. For each individual sample, 13 antibody functional properties against 8 *Mtb* and 4 non-*Mtb* microbial antigens were measured and analyzed via univariate and multivariate machine-learning approaches.

**Findings:** FcαR/CD89 immune complex formation with antibodies reactive to four *Mtb* antigens including ESAT-6 & CFP-10, FcγRI/CD64 associated with one *Mtb* antigen, and HIV gp120 IgA1 levels decreased during the intensive and continuation/consolidation phases of TB therapy. This antibody signature also highlighted treatment non-responsiveness and distinguished children with from those without TB disease with predictive capacity similar to Xpert.

**Interpretation:** An *Mtb* and HIV reactive peripheral blood antibody functional signature of FcαR/CD89, FcγRI/CD64, and IgA1 has the potential to complement current clinical tools and those in development to diagnose pulmonary TB disease in CLHIV.

**Funding:** This work is supported by UT Southwestern Disease Oriented Scholars Award (LLL), NIAID 5R01AI158858 (LLL), Burroughs-Wellcome Fund UTSW Training Resident Doctors as Innovators in Science (YJK), NICHD K12HD000850, NIAID K23AI143479 and R21AI192086 (LMC), NICHD R01 HD023412 (GJS), NIAID 75N93019C00071 (CW).

## Introduction

Tuberculosis (TB) is a leading cause of morbidity and mortality among children living with HIV (CLHIV)^1^. Even with antiretroviral therapy (ART) normalizing CD4 counts and suppressing viral replication, CLHIV compared to their uninfected peers remain at higher risk for acquiring TB and have worse outcomes^2^. Non-specific clinical symptoms, paucibacillary disease, delays in processing induced sputa, gastric aspirate and stool because of the inability to expectorate sputa, and limited performance of diagnostics on non-respiratory samples limit the ability to identify and treat TB disease in CLHIV^3^.

Current diagnostics developed and used in adults perform worse in children^4^. Culture of *Mycobacterium tuberculosis (Mtb),* the causative agent, is the gold standard and confirms 80-85% of adult^5,6^ but only 10-50% of pediatric^7^ cases. Smear for acid fast bacilli, PCR for DNA with Xpert Ultra, and urine lateral flow lipoarabinomannan (LF-LAM) in the HIV population are also more limited in children compared to adults^8–12^. As such, more than half of children with TB younger than 5 years of age are not diagnosed^1,13^.

Serological assays that detect levels of *Mtb* reactive antibodies are imprecise^14^, but antigen-specific antibody Fc receptor (FcR) engagement and subsequent immune cell effector functions are promising biomarkers of TB disease^15,16^. The inclusion of post-translational glycosylation that modulates FcR binding enhances the ability of *Mtb* reactive IgG to differentiate between classical pulmonary active TB disease and latent TB infection in HIV negative adults^17^. Moreover, *Mtb* reactive IgG glycosylation, FcR binding, and Fc effector functions in combination with subclasses are able to identify a spectrum of *Mtb* exposure and immune responses in adults not captured by current clinical diagnostics: HIV negative and positive close contacts of TB patients without a diagnosis of classical latent TB infection^18–20^, TB treatment response in HIV uninfected adults^21,22^, adolescents and possibly even adults who progress to TB disease^22,23^, and central nervous system disease^24^. The development of point-of-care devices to enhance operational capacity of a sample sparing microfluidic antibody functional platform shows the feasibility of leveraging antibody functional properties to complement current clinical diagnostic tools^25,26^. While IgG is the most well studied isotype, IgA and IgM can also modulate immune responses in TB^16,27–31^. How they may be biomarkers of disease particularly in children with an immature immune system and limited B cell responses are not clear^32^.

This study evaluated antigen-specific antibody functional properties in the serum of a clinically well-characterized cohort of Kenyan CLHIV with the goal of identifying a humoral immune signature of pulmonary TB disease^33^. 13 antibody functional properties (IgA1, IgM, IgG, IgG1, IgG2, IgG3, IgG4, immune complex formation with FcγRI/CD64, FcγRIIa/CD32a, FcγRIIb/CD32b, FcγRIIIa/CD16a, FcγRIIIb/CD32b, FcaR/CD89) against 12 microbial antigens (eight associated with *Mtb* and four non-*Mtb* pathogens including HIV) were measured.

FcαR/CD89 immune complex formation with antibodies reactive to four *Mtb* antigens including ESAT-6 & CFP-10, FcγRI/CD64 with one *Mtb* antigen, and HIV gp120 IgA1 levels could predict the presence of TB disease and responsiveness to TB therapy. These data suggest that peripheral blood antibody functional signatures could inform TB diagnostics and treatment monitoring in CLHIV.

## Results

### IgA, IgM and FcR immune complex formation decrease with effective TB treatment

To identify correlates of disease, we examined samples from 28 children collected longitudinally at three timepoints over the course of TB therapy: 1) pre-treatment, with the highest and most transmissible bacterial burden in active pulmonary TB disease, 2) intensive phase, with four anti-TB drugs rapidly killing actively replicating bacteria, and, 3) continuation or consolidation phase, with two anti-TB drugs focused on eliminating dormant bacteria to ensure cure and prevent relapse^4,34^ (Figure 1). The median age was 2.2 years and CD4% 13.5 before concomitant treatment for HIV and TB (Figure 1 and Supplemental Figure 1).

**Figure 1.**
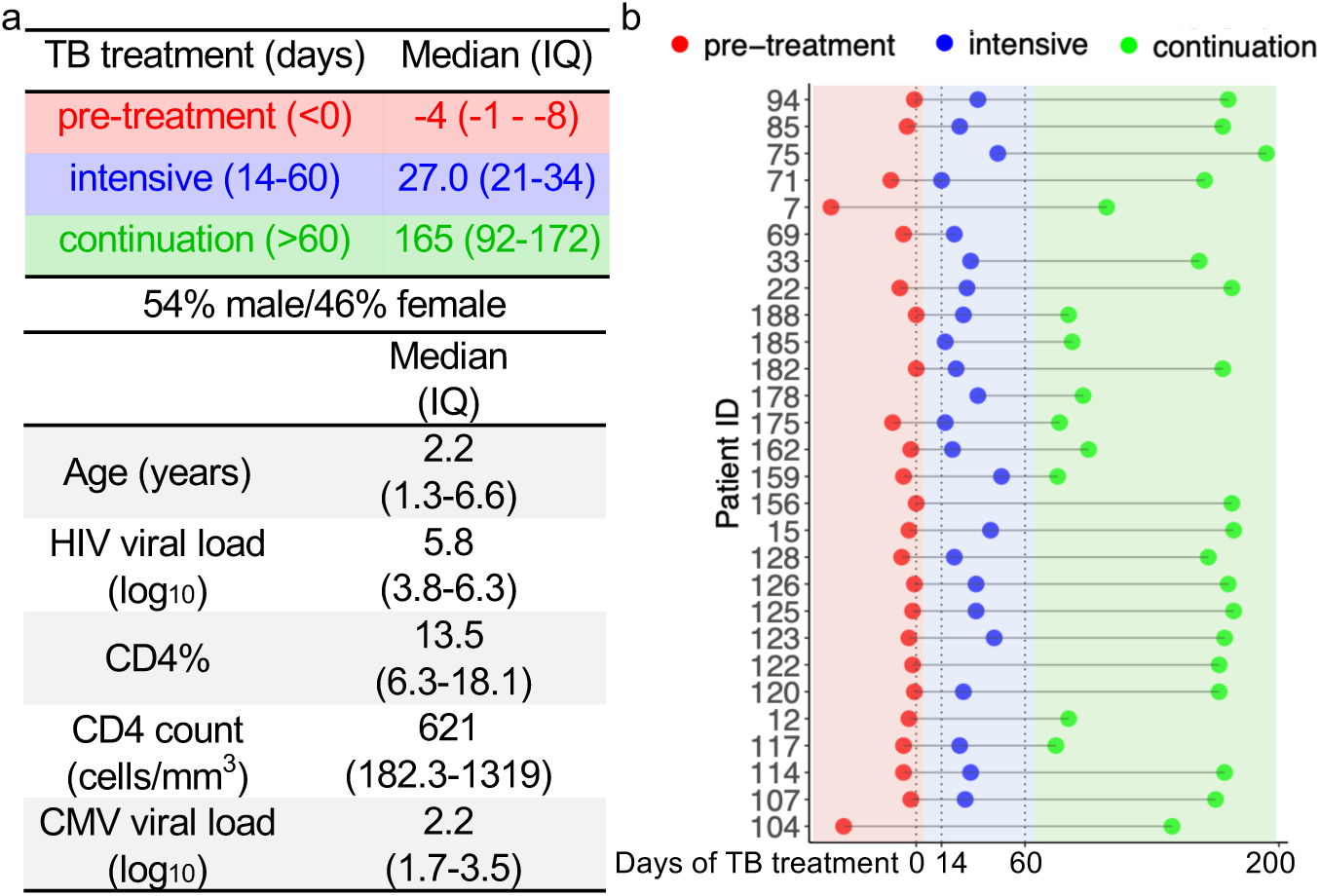
Samples collected longitudinally over the course of TB treatment in children living with HIV. Individuals were followed longitudinally before (pre-treatment) and during (intensive and continuation) TB treatment. Median and interquartile (IQ) ranges of sample collection days, age, HIV viral load, CD4%, CD4 counts and CMV viral load, along with sex distribution are shown (a). Each line represents a single individual and dots represent when samples were collected relative to TB treatment (b).

We measured antigen-specific antibody isotype, subclass, and Fc receptor binding using a customized, high-throughput, sample sparing Luminex approach^35^. Antibodies function via the Fab domain binding to microbial antigens in combination with the Fc domain binding to Fc receptors (FcRs) expressed on innate and adaptive immune cells^15^. The engagement of activating FcRs that bind to IgG (FcγRI/CD64, FcγRIIa/CD32a, FcγRIIIa/CD16a, FcγRIIIb/CD16b) and IgA (FcαR/CD89) in combination with inhibitory FcR (FcγRIIb/CD32b) regulate immune cell effector functions. All have been associated with different states of TB in human and animal models but how they reflect TB disease in CLHIV is not known^16,27,28,36–42^.

To identify *Mtb* reactive antibody responses, we used individual antigens associated with mycobacterial pathogenesis (Figure 2A): secreted virulence factors ESAT-6 & CFP-10 that are also used in current IFNγ-based diagnostics, Ag85A&B found in the Bacillus Calmette-Guérin (BCG) vaccine and also on the cell surface of and secreted from *Mtb*, the mycobacterial cell wall envelope lipomannan (LM) and mannose-capped lipoarabinomannan (ManLAM), and the periplasmic phosphate-binding lipoprotein PstS1. To capture the breadth of the more than 4000 open reading frames in *Mtb* representing antigens against which antibodies could potentially target^43,44^, we included preparations of proteins from *Mtb* bacterial culture with purified protein derivative (PPD) and whole cell lysate as well as the fraction secreted into the culture filtrate. To assess HIV reactive responses, we used the gp120 glycoprotein essential for infection (Figure 2B). Because cytomegalovirus (CMV) infection associates with both TB and HIV independently^45–47^, we used the major structural protein pp65 (Figure 2B). As control antigens reflective of common primarily pulmonary pathogens, we used influenza hemagglutinin and pneumococcal capsular polysaccharide mix from the Pneumovax23 vaccine (Figure 2B).

**Figure 2.**
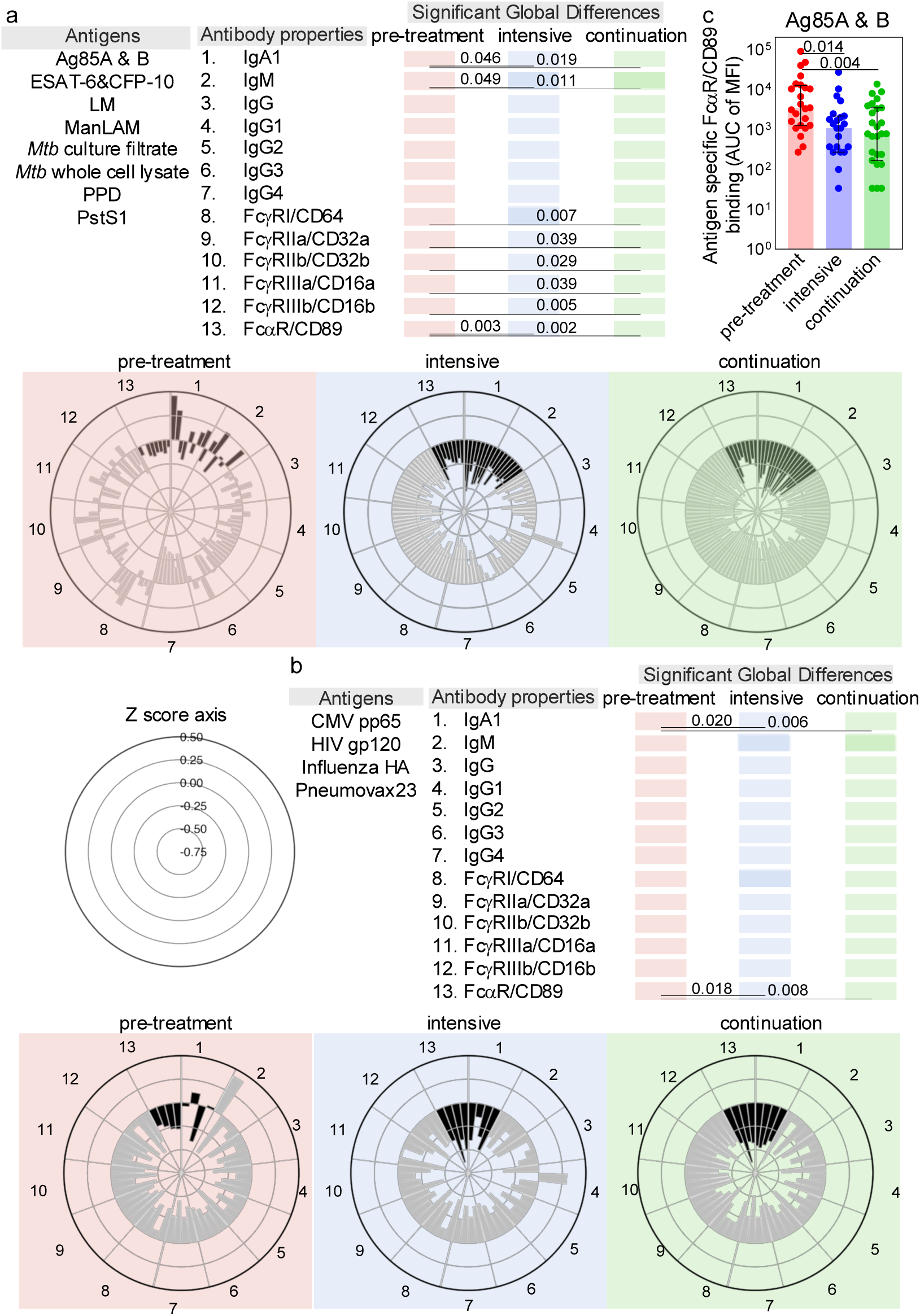
Evolution of antigen specific antibody isotypes, subclass, and Fc receptor immune complex formation with TB treatm ent. For each patient sample, relative levels of antigen specific antibodies and immune complex formation with Fc receptors (FcRs) were determined by customized multiplex Luminex. Area under the curve (AUC) summarizes median fluorescence intensities (MFIs) from sample serial dilutions. Petal plots show the Z-scored data for each antigen specific antibody isotype and subclass and FcR immune complex formation across samples collected longitudinally from individuals before (red, N=24) and during intensive (blue, N=23) and continuation (green, N=27) phases of TB treatment. Partial Mann-Whitney p values combined by Fisher’s method summarize significance of changes within each isotype, subclass, and FcR for antibodies reactive to *Mtb* (a) and non-*Mtb* (b) antigens. Bars depict the medians and interquartile ranges for A85A & B FcαR/CD89 immune complex formation with each dot representing one single individual patient sample. Kruskal-Wallis determined significance (c).

In general, we found that *Mtb* reactive antibodies and FcR immune complex formation were highest in TB disease before treatment and decreased over the course of intensive and continuation phases (Figure 2). Globally, *Mtb* reactive IgA1, IgM, and immune complex formation with FcαR/CD89 decreased in the first two months of intensive TB treatment and in the consolidation/continuation phase (Figure 2A and 2C). *Mtb* reactive IgG immune complex formation with FcγR, not levels of the isotype or its subclasses, were during the consolidation/continuation phase (Figure 2A). For non-*Mtb* antigens, IgA1 and immune complex formation with FcαR/CD89, not IgM, IgG, or FcγR decreased (Figure 2B). These data suggest that generalized hypergammaglobulinemia in IgA1 and the functional ability to bind to FcαR/CD89 characterizes TB disease in CLHIV and rapidly responds to treatment (Figure 2).

One child in the group was clinically non-responsive to TB treatment^48^. After 4 months of TB therapy, fevers, chills, cough, and weight loss persisted. To assess if the pre-TB treatment humoral immune signature could capture this clinical outlier, we used sparse principal component analysis to incorporate the 13 antibody properties measured for each of the 12 microbial antigens (eight *Mtb*, one influenza, one CMV, one pneumococcal, and one HIV) (Supplemental Figure 2A and 2B). We observed that IgA1 and FcαR/CD89 immune complex formation involving *Mtb* antigens represent the top five antibody properties that distinguished this non-responder from responders (Supplemental Figure 2B and 2C). These analyses show that the pre-TB treatment antibody signature that predicts clinical non-responsiveness to TB treatment (Supplemental Figure 2) aligns with that which distinguishes disease before effective treatment (Figure 2). Thus, an FcαR/CD89 immune complex formation and IgA1 has the potential to predict response to TB therapy.

### TB therapy associates with progressively coordinated decreases in FcR binding, IgA and IgG

We next used a nested mixed linear model to assess antibodies at the individual level of each child for whom paired samples were collected at pre-treatment and intensive (Figure 1B and 3A) as well as pre-treatment and continuation/consolidation (Figure 1B and 3B) time-points. Within the early timeframe of the intensive phase of treatment, 29 antibody properties and functions decreased (Figure 3A). This number increased to 95 at the consolidation/continuation timepoint marking longer treatment duration (Figure 3B). The 29 *Mtb* and non-*Mtb* antibody properties that responded early in treatment were characterized by 10 antigen-specific IgA1, eight involving FcαR/CD89, six involving low affinity FcγRs (primarily FcγRIIIb/CD16b), three involving the high affinity FcγRI/CD64, two antigen-specific IgM, and no IgG or its subclasses (Figure 3A and Supplemental Table 1). Along with these 29, an additional 66 antibody properties decreased by the consolidation/continuation time-point with 34 involving FcγR, 24 IgG and subclasses, five IgM, and three IgA1 (Figure 3B and Supplemental Figure 2). These results on the level of each individual child (Figure 3) were consistent with those at a more global level (Figure 2) in showing that IgA1 and FcαR/CD89 as well as some IgM respond rapidly and persistently to TB treatment whereas changes in IgG and FcγR immune complex formation are delayed.

**Figure 3.**
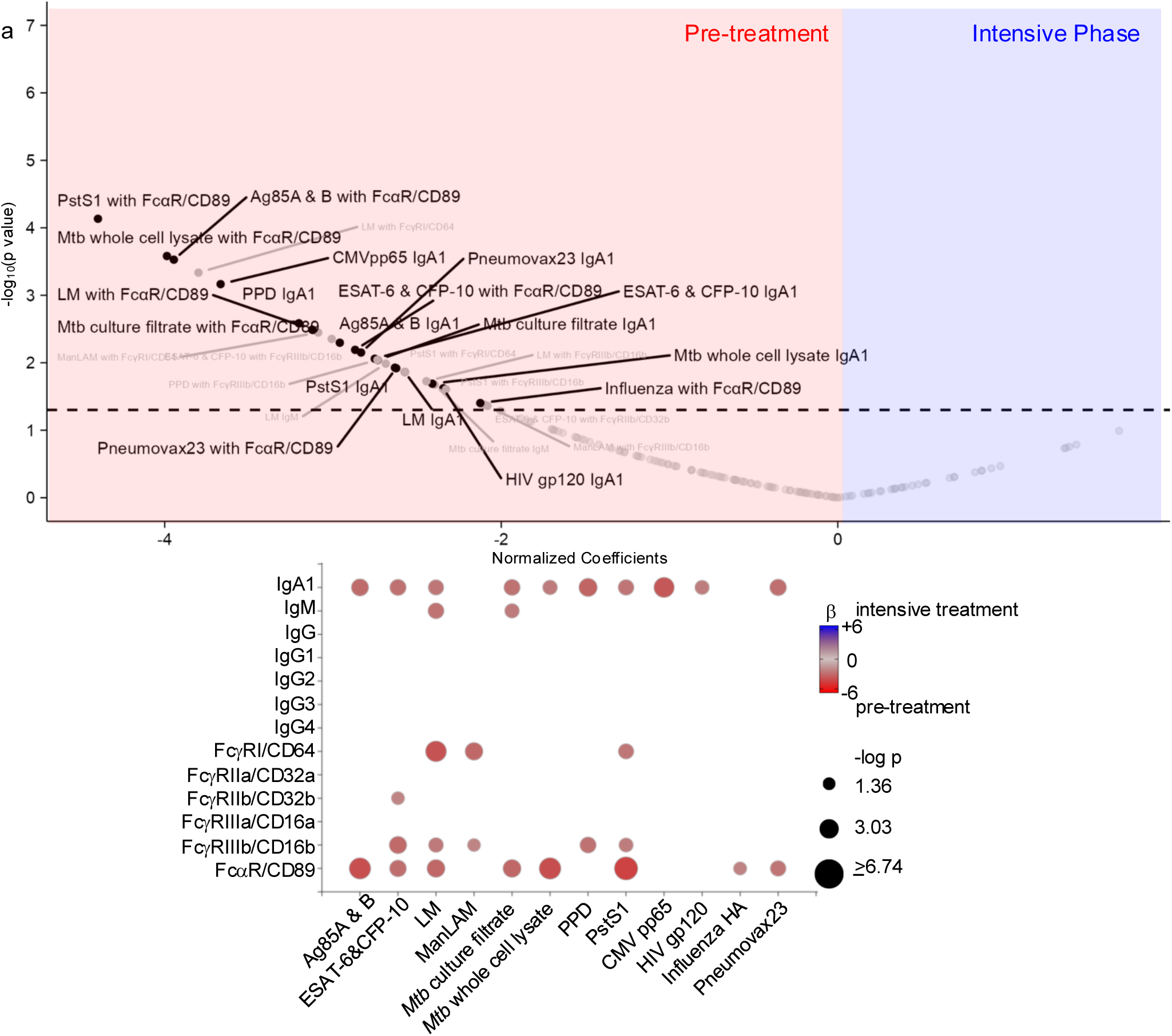
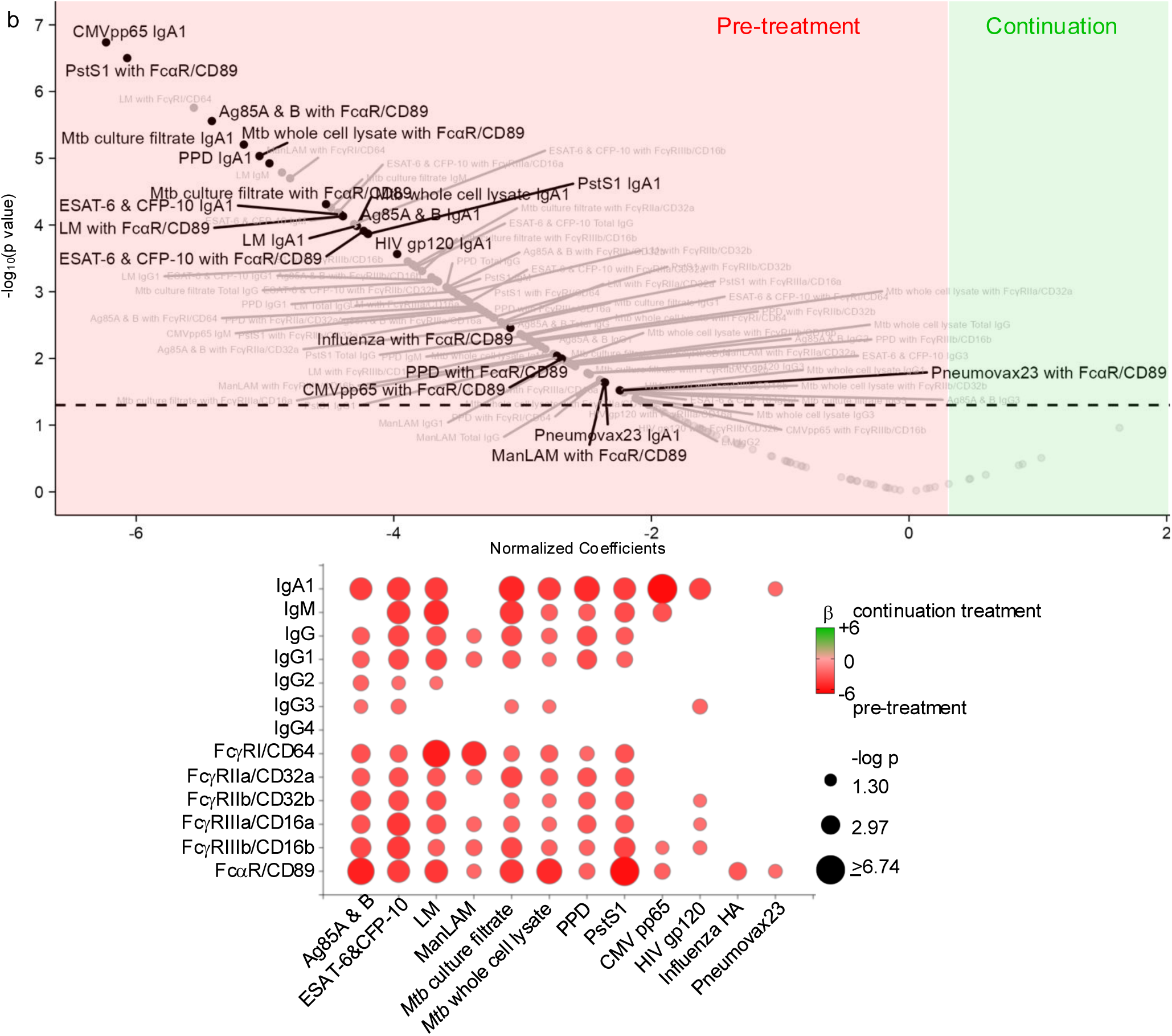
Effects of TB treatment on antigen specific antibody Fc properties in children living with HIV. Volcano plots show normalized coefficients for 156 antigen specific antibody properties and functions highlighted in pre-treatment (red, N=23) compared to intensive (blue, N=22) (a) and continuation (green, N=26) (b) treatment phases from mixed linear regression adjusting for pre-enrollment age, sex, CD4%, HIV viral load. IgA1 and FcαR/CD89 properties are highlighted. Dotted line marks p=0.05. Normalized coefficients for antigen specific antibody properties p≤0.05 are depicted in bubble plots.

From the many antigen-specific antibody properties and functions that decrease with TB treatment, we sought to identify those that most characterized changes. Using LASSO (Least Absolute Shrinkage and Selection Operator) to select the minimum number of features and PLS-DA (Partial Least Squares Discriminant Analysis), we identified from the initial N=156 antigen-specific antibody properties a subset discriminating pre-treatment from intensive (Figure 4A) and pre-treatment from continuation/consolidation (Figure 4B) time-points. Immune complex formation between FcαR/CD89 and antibodies recognizing the *Mtb* associated Ag85A&B,

**Figure 4.**
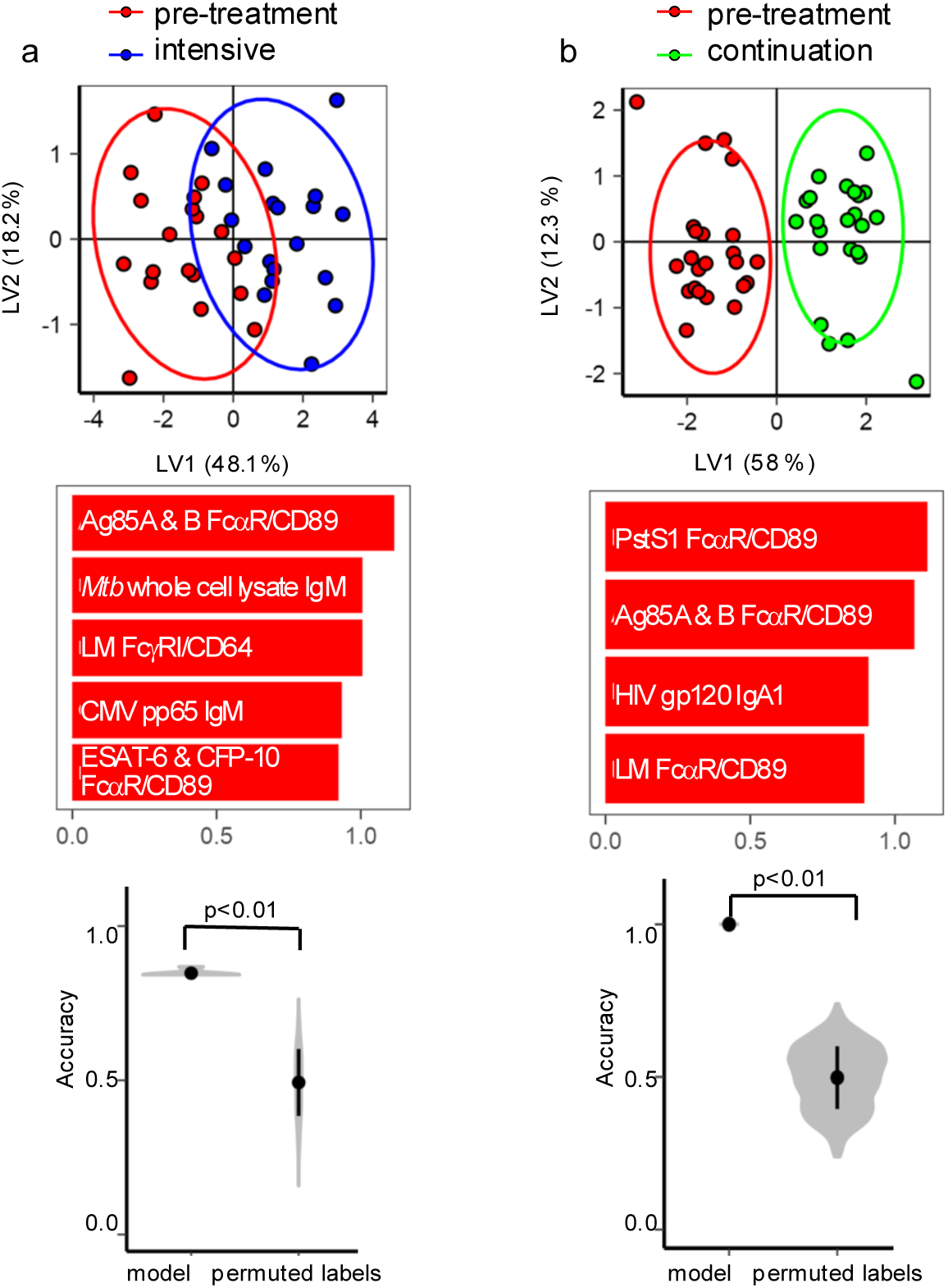
Selected antigen specific antibody biomarkers that distinguish phases of TB treatment in children living with HIV. PL SDA with LASSO using data from N=28 individuals was used to select antigen specific antibody properties and functions that separated pre-treatment (red) from intensive (blue) (a) and continuation (green) (b) TB treatment phase groups. Ellipses mark 95% confidence interval s. Minimal number of features to separate groups are shown in Variable Importance in Projection (VIP) loadings plots. Performance was assessed by 5 fold cross validation. Violin distribution plots show median classification accuracies using selected (model, n=10) compared to random permutations (permuted labels, n=1000).

ESAT-6 & CFP-10, PstS1, and LM were highlighted in addition to *Mtb* whole cell lysate and CMV pp65 IgM, HIV gp120 IgA1, and immune complex formation between LM IgG and FcγRI/CD64. Co-correlates network analyses of LASSO-selected antibody properties demonstrated clustering with *Mtb* and non-*Mtb* reactive antibody properties in the intensive (Supplemental Figure 5A) and even more so the consolidation/continuation timepoint (Supplemental Figure 5B). These analyses demonstrate the progressive and collaborative nature of polyclonal responses where changes occur in the context of many antigen-specific antibodies rather than in isolation over the course of TB treatment.

### A signature highlighted by FcαR/CD89 and IgA predicts TB disease in children living with HIV

To validate the antigen-specific antibody properties identified as biomarkers of TB disease following TB treatment, we used an orthogonal cross-sectional approach comparing CHLIV with untreated TB disease to those with no TB disease (Figure 5). We identified by multiple linear regression 27 *Mtb* and non-*Mtb* reactive antibody properties and functions higher in children with compared to without TB disease (Figure 6). 12 were IgA1, eight involved FcαR/CD89, three involved FcγRIIIb/CD16b, two were IgG2, one involved FcγRI/CD64, and one was IgG3 (Supplemental Table 3). Adjustments for age, sex, CD4% and HIV viral load at enrollment were made given variable relationships with antibody properties (Supplemental Figure 4). No antigen-specific antibody property or function was lower in those with compared to without TB disease (Figure 6).

**Figure 5.**
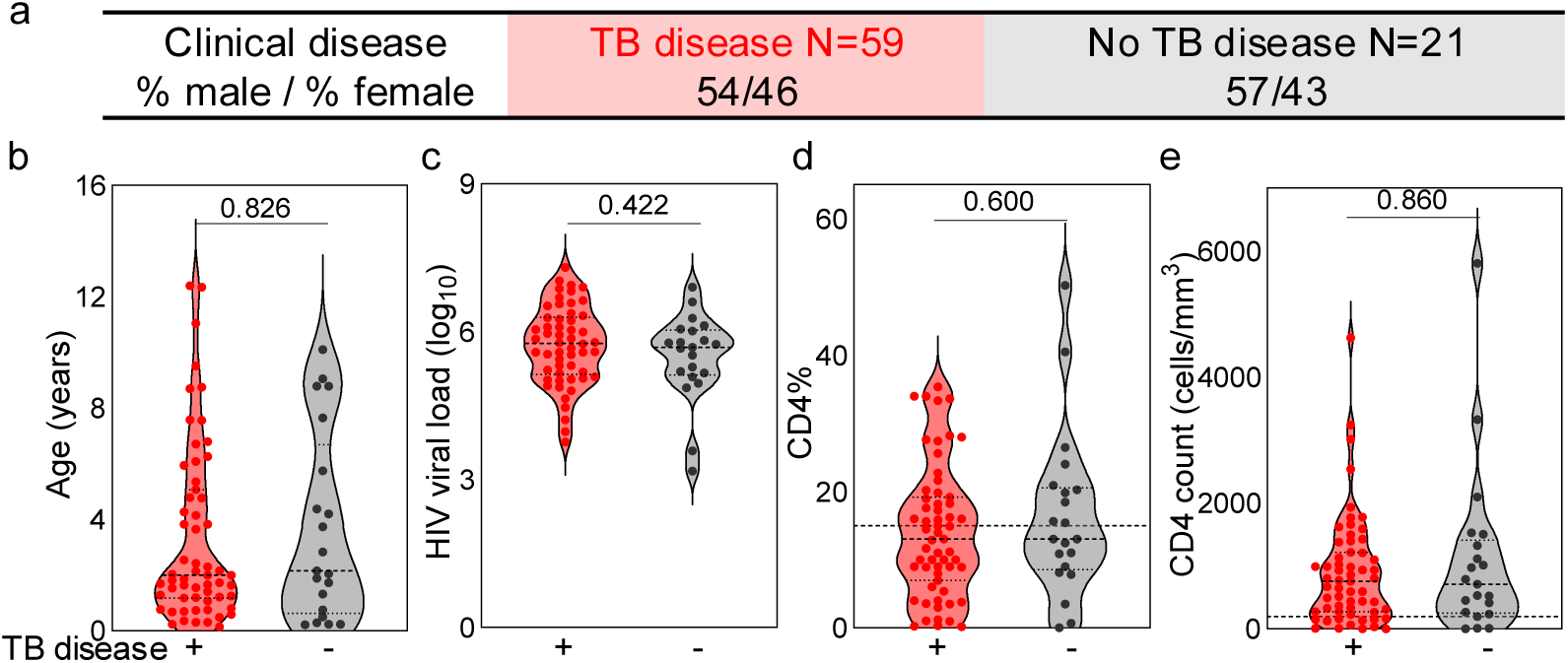
Samples collected from children living with HIV with and without untreated TB disease. Number and distribution of sex are shown for individuals with and without TB disease (a). Violin plots show median and range of age (b), HIV viral load (c), CD4% (d), and CD4 count (e). Dotted lines mark CD4%=15 and CD4 absolute count=200. Mann-Whitney U determined significance.

**Figure 6.**
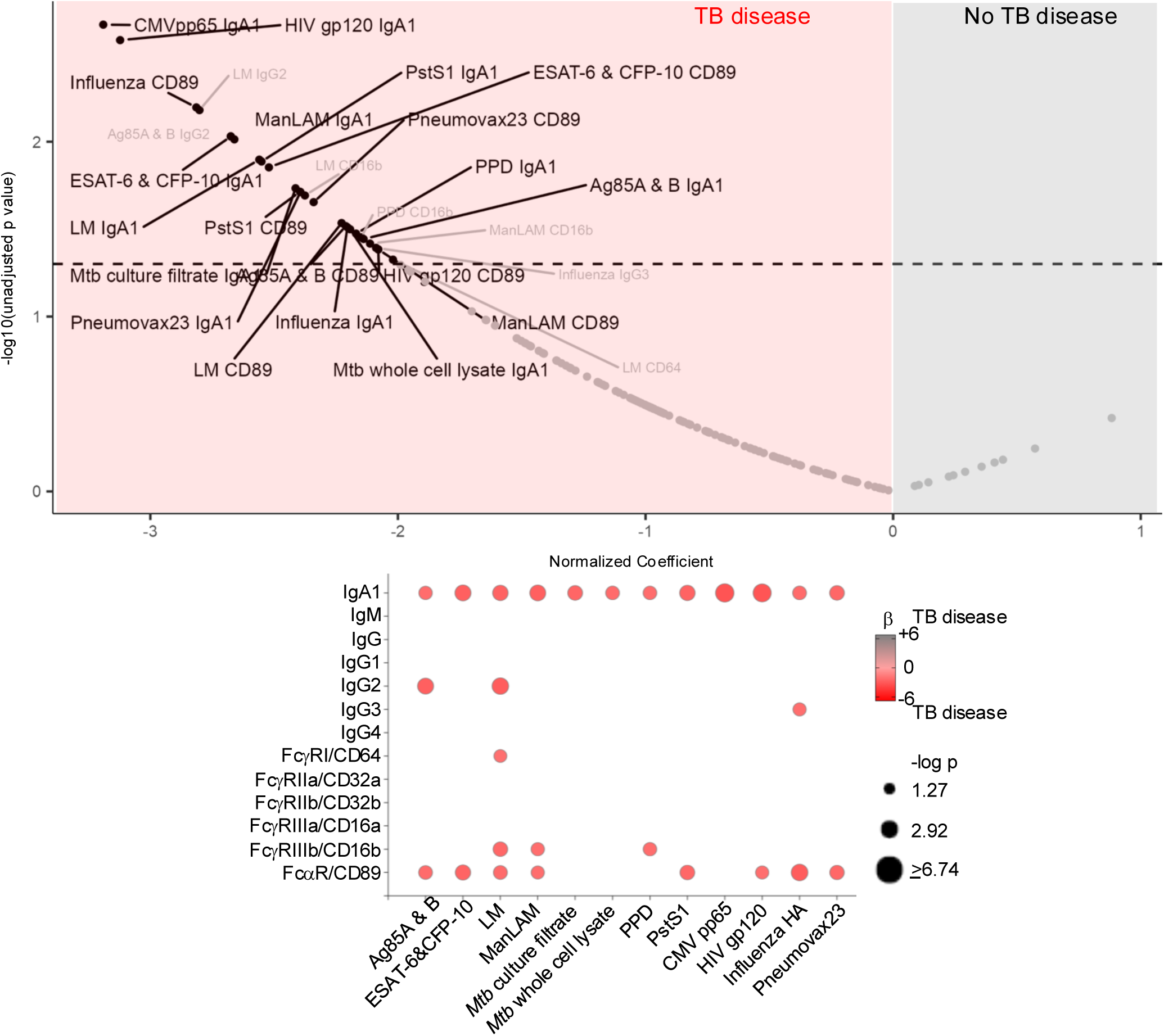
Antigen specific antibody Fc properties in children with compared to children without TB disease. Volcano plot shows normalized coefficients for 156 antigen specific antibody properties and functions highlighted in individu als with (red, N=59 individuals) compared to without (grey, N=21 individuals) TB disease from multiple linear regression adjusting for pre-enrollment age, sex, CD4%, HIV viral load. IgA1 and FcαR/CD89 properties are highlighted. Dotted line marks p=0.05. Normalized coefficients for antigen specific antibody properties p≤0.05 are depicted in bubble plots.

To identify antigen-specific antibody properties and functions that could both mark the presence of TB disease across different children and change with treatment within each child, we cross referenced results from cross-sectional (Figure 6) and longitudinal approaches (Figure 3). We found that 20 were identified in both analyses (Figure 7A). 10 represented IgA1, six involved FcαR/CD89, three involved FcγRIIIb/CD16b, and one involved FcγRI/CD64. Of the 20, six overlapped with those selected by PLSDA with LASSO (Figure 4 and 7A-7G). Receiver operator curve (ROC) analyses showed a range of areas under the curves (AUCs) from 0.67 to 0.74 (Figure 7H-M). Incorporating all six into a single model resulted in an AUC with a mean value of 0.76 (Figure 7N). Together, these data show that the abilities of five *Mtb* reactive antibodies to immune complex with FcαR/CD89 and FcγRI/CD64 along with HIV IgA1 can predict the presence of untreated TB disease in CLHIV.

**Figure 7.**
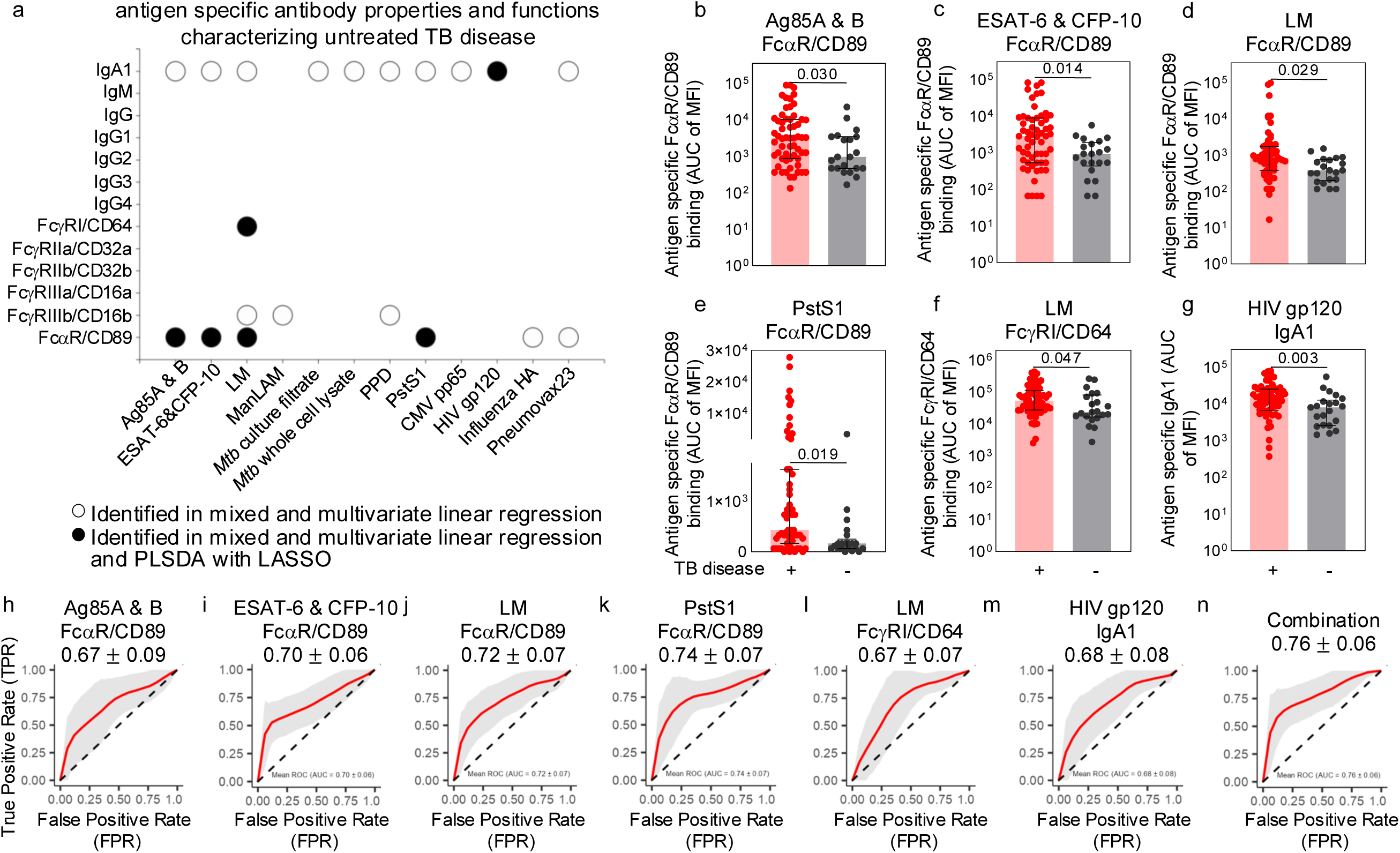
Predictive capacity of antigen specific antibody properties for TB disease in children living with HIV. Bubble plot shows antigen specific antibody properties and functions identified from both longitudinal and cross-sectional data analyzed by mixed and multiple linear regression models (all circles) as well as PLSDA with LASSO (black circles) (a). Bars in column plots depict medians and interquartile ranges of the six highlighted in all analyses that identify the presence of TB disease (black circles in a) in individuals with (red) and without (grey) TB disease (b-g). Significance was calculated by multiple linear regression adjusting for age, sex, and baseline CD4% and HIV viral load. Receiver operator curves (ROC) demonstrate the ability to predict the presence of untreated TB disease for the same six individually (H-M) and in combination (N). Grey areas show min and max from 100 iterations of random sampling with estimated mean depicted by the red line and denoted along with standard error of the mean.

## Discussion

This study evaluated the potential of antibody functional properties to identify TB disease in a clinically well characterized cohort of CLHIV^33^. Two orthogonal approaches were utilized.

The first longitudinally followed children over the course of TB treatment (Figure 1). The second assessed a cross section of children with and without untreated TB disease (Figure 5). We found that FcαR/CD89 immune complex formation, IgA1, and some IgM began to decrease in the first two months of intensive TB treatment (Figures 2, 3A). FcγR immune complex formation and IgG followed in the continuation/consolidation phase of TB treatment (Figure 2, 3B).

FcαR/CD89 immune complex formation and IgA1 distinguished one child who did not respond to TB treatment from those who clinically improved, suggesting that this humoral immune signature reflected changes in disease and not anti-TB therapy itself (Supplemental Figure 2). PLSDA with LASSO selected FcαR/CD89 immune complex formation to separate pre-treatment from intensive (Figure 4A) as well as consolidation/continuation phases of TB treatment (Figure 4B). Independent orthogonal cross-sectional evaluation identified an enhanced antibody signature dominated by IgA1 and immune complex formation with FcαR/CD89 in HIV infected children with compared to without untreated TB disease (Figure 6). Four of the six antigen-specific antibody properties and functions identified from both longitudinal and cross-sectional data involved FcαR/CD89 complex formation with *Mtb* antigens including ESAT-6 & CFP10 (Figure 7A-E). The other two were the *Mtb* reactive LM FcγRI/CD64 and HIV gp120 IgA1 (Figure 7A, F, G). AUCs of ROCs for these six antigen-specific antibody properties and functions from 0.67 to 0.74 individually and 0.76 in combination (Figure 7H-N) demonstrated the potential to predict untreated TB disease in CLHIV.

Previous studies show that *Mtb* reactive IgA associate with protective and pathologic consequences via FcαR/CD89 dependent and independent mechanisms. In mice, the presence of human FcαR/CD89 is necessary for pulmonary delivered IgA to protect against *Mtb* aerosol infection^27,28^. In human lung epithelial A549 cells, FcαR/CD89 expression is not necessary for monoclonal and polyclonal IgA from adults with untreated pulmonary TB to inhibit *Mtb* uptake^29^. In non-human primates, pre-*Mtb* infection levels of arabinomannan reactive IgA in bronchoalveolar lavage associate with the absence of TB disease developing after aerosol inoculation^30^. In contrast to potential protection from pulmonary IgA, *Mtb* reactive IgA in the peripheral blood of HIV negative adults^49,50^ and adolescents adolescents^23^ variably correlate with progression to disease. The association between *Mtb* reactive IgA and immune complex formation with FcαR/CD89 and pulmonary disease is more strongly observed in a cross-section of HIV negative children^51^. Together, these data suggest that an association between pulmonary TB disease and FcαR/CD89 immune complex formation by peripheral blood antibodies is independent of HIV co-infection and geographical region.

Antigen-specific antibody functional properties have the potential to enhance current diagnostics in the detection of transmissible *Mtb* from pulmonary disease. We and others have shown that IgG post-translational glycosylation leading to differential FcγR binding and Fc effector functions have equivalent if not better potential than levels of antigen-specific antibody subclasses to identify TB disease in HIV negative and positive adults^17,18,21,22,40,52^. Consistent with these data, expression of the high affinity FcγRI/CD64 in peripheral blood has been associated with TB disease across species in the absence and presence of HIV^42,53^. In this study both levels of antigen-specific IgA1 and immune complex formation with FcαR/CD89 were measured but only the latter was selected consistently by analyses of both longitudinal and cross-sectional data to associate with TB disease (Figure 4, 7A-E and H-K). In addition, LM FcγRI/CD64 correlated with disease more than LM IgG and subclasses (Figure 3, Figure 7A, F, L). These results emphasize the consequences of *Mtb* reactive antibody immune cell effector functions rather than antibodies in isolation. The data suggest that antibody induced phagocytosis, reactive oxygen species, antigen presentation, cellular cytotoxicity and cytokine release in neutrophils, monocytes, macrophages, and dendritic cells via FcαR/CD89 and FcγRI/CD64 can modulate *Mtb*^54^.

In comparison to microbial biomarkers, those that are host-based can be affected the immune substrate. Global hypergammaglobulinemia and the high burden of microbial pathogens in severe immunosuppression associated with untreated HIV, as well as interactions between HIV, CMV, and TB^45,47^ could explain how both *Mtb* and non-*Mtb* reactive antibody properties and functions highlight the presence of TB disease (Figures 2-4 and 6-7). Specifically, the ability of HIV gp120 IgA1 to predict TB disease likely reflects the high burden of HIV in these children and impact of HIV on humoral immunity in TB (Figure 7)^52,55,56^.

There are several inherent limitations to this study. First, the results relate to pulmonary disease, not disseminated or extrapulmonary forms of TB that have been observed in younger children. Second, the data reflect Kenyans and not necessarily children in other geographical regions with potentially different endemic mycobacterial exposure. Third, while IgA, IgG, and IgM cross from the blood into the lung, what can be extrapolated from these data as to pulmonary interactions between *Mtb* and the host humoral immune response is indirect.

As peripheral blood biomarkers, these data show that antibody functional properties have the potential to fill the significant need for improved TB diagnostics in children living with HIV. Consistent with other studies, sensitivity and specificity for WHO recommended diagnostics such as Xpert on stool (60% sensitivity, 98% specificity) is equivalent to sputum (63% sensitivity, 99% specificity) and higher than urine LAM (43% sensitivity, 91% specificity) for children in this cohort^57,58^. The data here show that the capacity for *Mtb* reactive antibodies to immune complex with FcαR/CD89 and FcγRI/CD64 in combination with HIV reactive IgA1 to predict TB disease (Figure 7N) may be equivalent to Xpert in meta-analyses^12^ and other host-based biomarkers such as the monocyte-lymphocyte ratio (MLR) (AUC 0.74) and even better than urine LAM (AUC 0.67)^59,60^. Thus, an *Mtb* and HIV peripheral blood antibody functional signature has the potential to enhance current clinical tools and those in development to identify CLHIV who would benefit from treatment for TB disease^61–66^.

## Materials and Methods

### Patient TB samples

This study is nested within the Pediatric Urgent Start of HAART” (PUSH) clinical trial in Kenya (ClinicalTrials.gov: NCT02063880)^33^. Between April 2013 and November 2015, 181 children under 13 years of age who were diagnosed with HIV at hospital admission and antiretroviral therapy (ART)-naive were enrolled and randomized to start ART within 48 hours (urgent) versus 7-14 days (early). Children with extrapulmonary TB disease such as central nervous system (CNS) involvement and other active opportunistic infections or chronic conditions known to affect the immune system were excluded from these studies. TB disease was assessed by clinical signs and symptoms of disease, chest x-ray, two sputa or gastric aspirate samples for mycobacterial culture and Xpert *MTB*/RIF (Cepheid, USA), and one stool Xpert *MTB*/RIF. Because heterogeneity of pediatric TB clinical diagnosis is an inherent challenge to this field of study, TB classification was made based on international consensus definitions for pediatric TB^48^. TB treatment was initiated per National Kenyan Guidelines^67^ with rifampicin, isoniazid, pyrazinamide, and ethambutol for the first 2 months (intensive phase) followed by rifampicin and isoniazid for the next 4 months (consolidation/continuation phase). Response to TB treatment was determined by symptom resolution and weight gain at the 6-month study follow-up visit.

Samples from 83 children were used in this study: n=6 pulmonary TB disease confirmed by culture or Xpert, n=56 clinically suspected/microbiologically unconfirmed pulmonary TB disease based on clinical symptoms and signs, radiographic, immunologic and treatment response, and n=21 with unlikely TB disease based on the absence of aforementioned criteria. Inclusion for this sub-study was based on TB disease classification and sample availability at enrollment and through TB treatment follow-up. Groups were balanced by age, sex, baseline CD4%, and HIV viral load (VL). 97% (n=60) of children were BCG vaccinated in the TB disease group and 100% (n=21) in the unlikely TB disease group. Urgent compared to post-stabilization ART was initiated in 53% of children with TB disease and 43% of children without TB disease. Blood samples were collected in sodium heparin tubes, centrifuged at 1500 rpm, sera removed, and stored at -80°C. Written informed consent was obtained from the legal primary caregivers of enrolled children. The study was approved by the University of Nairobi Ethics and Research Committee/Kenyatta National Hospital, and institutional review boards (IRBs) of the University of Washington, Emory University, and UT Southwestern Medical Center.

### Antigen-specific antibody isotype and subclass

Antigen-specific isotypes and subclasses were quantified as described with modifications^68,69^. Carboxylated microsphere beads (Luminex) were coupled to protein antigens by covalent NHS-ester linkages via 1-Ethyl-3-[3-dimethylaminopropyl]carbodiimide hydrochloride (EDC) and (N-hydroxysulfosuccinimide) (NHS) (Thermo Scientific) per manufacturer instructions and glycan antigens by 4-(4,6-dimethoxy[1,3,5]triazin-2-yl)-4-methyl-morpholinium (DMTMM)^70^. Antigen sources were as follows: PstS1 (BEI Resources NR-14859), Ag85A and B (BEI Resources NR-14871, NR-14870), ESAT-6 and CFP-10 (BEI Resources NR-49424, NR-49425), PPD (Statens Serum Institute), H37Rv whole cell lysate (BEI Resources NR-14822), H37Rv culture filtrate (Resources BEI NR-14825), HIV Clade A GP120 (Abnova gp120 92RW020), CMV pp65 (Abcam, ab43041), influenza hemagglutinin (BEI Resources NR-51702, NR-51401), mannosylated lipoarabinomannan (ManLAM) (BEI Resources NR-14848), Lipomannan (LM) (BEI Resources BR-14850) and pneumococcal polysaccharide (Pneumovax23 from Merck & CO., INC). Antigen coupled microspheres (1000 beads per well) were incubated with diluted serum (for IgG, IgG1 and IgG2, 1:300, 1:900, 1:2700 and for IgM, IgA1, IgG3 and IgG4, 1:100, 1:300, 1:900) in 96-well Bioplex Pro Flat Bottom plates (Bio-Rad) at 4°C for 16 hrs. After washing away the unbound antibodies, bead bound antigen-specific antibodies were detected by using a PE-coupled detection antibody for each isotype and subclass (anti-IgG, IgA1, IgM, IgG1, IgG2, IgG3 and IgG4 from Southern Biotech). After 2 hrs of incubation at room temperature, the beads were washed with PBS with 0.05% Tween-20 and fluorescence intensity acquired on a MAGPIX instrument containing xPONENT4.2 software (Luminex). For each individual sample, the relative level of antigen-specific antibodies was defined as the area under the curve (AUC) of serial dilutions.

### Antigen-specific antibody Fc receptor immune complex formation

Antigen-specific Fc receptor (FcR) immune complex formation was assessed as described with modifications^35,68,69,71^. Carboxylated microsphere beads (Luminex) were coupled as described above. Antigen coupled microspheres (1000 beads per well) were incubated with diluted sera (for FcγRIIIa/CD16a, FcγRIIa/CD32a, and FcγRIIb/CD32b 1:300, 1:900, 1:2700, for FcγRI/CD64, FcγRIIIb/CD16b, and FcαR/CD89 1:100, 1:300, 1:900) in 96-well Bioplex Pro Flat Bottom plates (Bio-Rad) at 4°C for 16 hrs. FcRs (FcγRIIIa/CD16a, FcγRIIIb/CD16b, FcγRIIa/CD32a, FcγRIIb/CD32b, and FcαR/CD89) (R&D) were conjugated with phycoerythrin (PE) (Abcam) per manufacturer’s instructions. FcγRI/CD64 (R&D) was added to IgG coated beads and then incubated with mouse anti-human FcγRI/CD64 (clone 10.1, Santa Cruz) (1 hr, room temperature) followed by PE-conjugated goat anti-mouse (Southern Biotech) (1 hr, room temperature). Fluorescence intensity was acquired on a Magpix instrument (Luminex), and AUC of serial dilutions calculated for each individual sample.

### Statistical Analyses

Medians and interquartile ranges summarize data (Figure 1A, 2A, 5B-E, 7B-G). Kruskal-Wallis (Figure 2C), Mann-Whitney U (Figure 5B-E), linear regression adjusting for age, sex and baseline CD4% and HIV viral load (Figure 7B-G), and paired Student t-test (Supplemental Figure 1) determined significance.

To summarize differences by isotype, subclass, and FcR (Figure 2A-B), partial Mann-Whitney U p-values for each antigen-specific antibody property were aggregated by Fisher’s method^72,73^ to compute a global test statistic and compared to a null distribution of 1,000 permutations to determine significance using the R package *rstatix*.

Mixed and multiple linear regression^74^ with the lme() function from the *nlme* package (Version 3.1.166) was used to identify differences in antigen-specific antibody properties (Figure 3 and 6), adjusting for age, sex, and baseline CD4% and HIV viral load at enrollment (Figure 6).

To identify pre-TB treatment antibody properties that distinguish a drug non-responder from responders, Z-scored antibody data was clustered by unsupervised Euclidean distance (Supplemental Figure 2A). Sparse Principal Component Analysis (Sparse PCA)^75,76^ with the optimal sparsity parameter selected through 5-fold cross-validation was used to identify the most influential features using the SPC() function from the *PMA* package (Version 1.2.4) (Supplemental Figure 2B-C).

Partial least squares discriminant analysis (PLS-DA)^77^ (Figure 4) was performed using the train_ropls() function from the systemsseRology package (Version 1.1)^78,79^, which interfaces with the ropls package (Version 1.36.0). Features selected by LASSO^80^ in more than a specified proportion of the 20 trials were retained for modeling. 10 five-fold cross validations assessed performance and p-values were calculated empirically as the proportion of permuted accuracies ≥ observed accuracy from 10 repetitions of 100 permuted label sets. For network analyses (Supplemental Figure 3), Spearman correlation coefficients were computed using the cor() function in base R (v4.3.2). The enrichment for each node was determined by comparing the group means, assigning the enrichment label to the group with the higher mean.

Receiver operating characteristic (ROC) (Figure 7H-N) was assessed using pROC package (v1.18.5)^81^. 100 bootstrap iterations were performed, with each randomly selecting 80% of the data while preserving the original class distribution. For each bootstrap sample, true positive rates were interpolated at 100 evenly spaced false positive rate points from 0 to 1. The mean ROC curve and 95% confidence interval were calculated across bootstraps. Mean and standard deviation are reported for AUC.

Data were visualized using Graphpad Prism 10, Excel 2016, *ggplot2* (Version 3.5.2), *ggrepel* (Version 0.9.6), *dplyr* (Version 1.1.4), *reshape2* (Version 1.4.4), *ggpubr* (Version 0.6.0), *tidyr* (Version 1.3.1), *pheatmap* (Version 1.0.12), igraph (version 2.0.3) and ggraph (version 2.2.1).

## Author contributions

YK designed the research study, conducted experiments, acquired and analyzed data, and wrote the manuscript. LMC designed the research study and experiments, provided the clinical samples, and wrote the manuscript. AM and NW analyzed data and wrote the manuscript. PL conducted experiments, acquired data, analyzed data, and wrote the manuscript. DW, GJS provided the clinical samples and IN, EM, SL, JS, GJS, DW provided feedback on the manuscript. CW analyzed the data and wrote the manuscript. LL designed the research study and experiments, conducted experiments, acquired and analyzed data, and wrote and revised the manuscript.

## Data sharing statement

All raw antibody data generated in this study, as well as metadata for all subjects, are included in a supplementary data file.

## Declaration of interests

Amyn A. Malik was employed by Analysis Group, Inc from 2022-2024.

## Data Availability

All data produced in the present study are available upon reasonable request to the authors

## Acknowledgements

We thank Joshua Miles, Gabrielle Lessen, and Nowrosh Islam for assistance with manuscript editing, Stephen Ostermann and Frederick Scott for technical support, study participants, caregivers, and the research administrative, clinical, and data teams for their dedication and support.

---

**Supplemental Figure 1.**
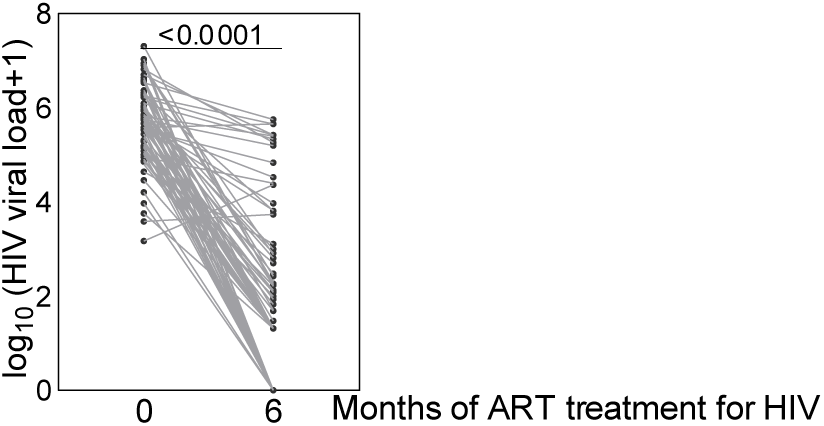
HIV viral load before and during antiretroviral (ART) treatment for HIV. Dot plot shows available HIV viral load at 0 and 6 months post initiation of ART treatment for individuals (N=78). Paired Students’s t-test determined significance.

**Supplemental Figure 2.**
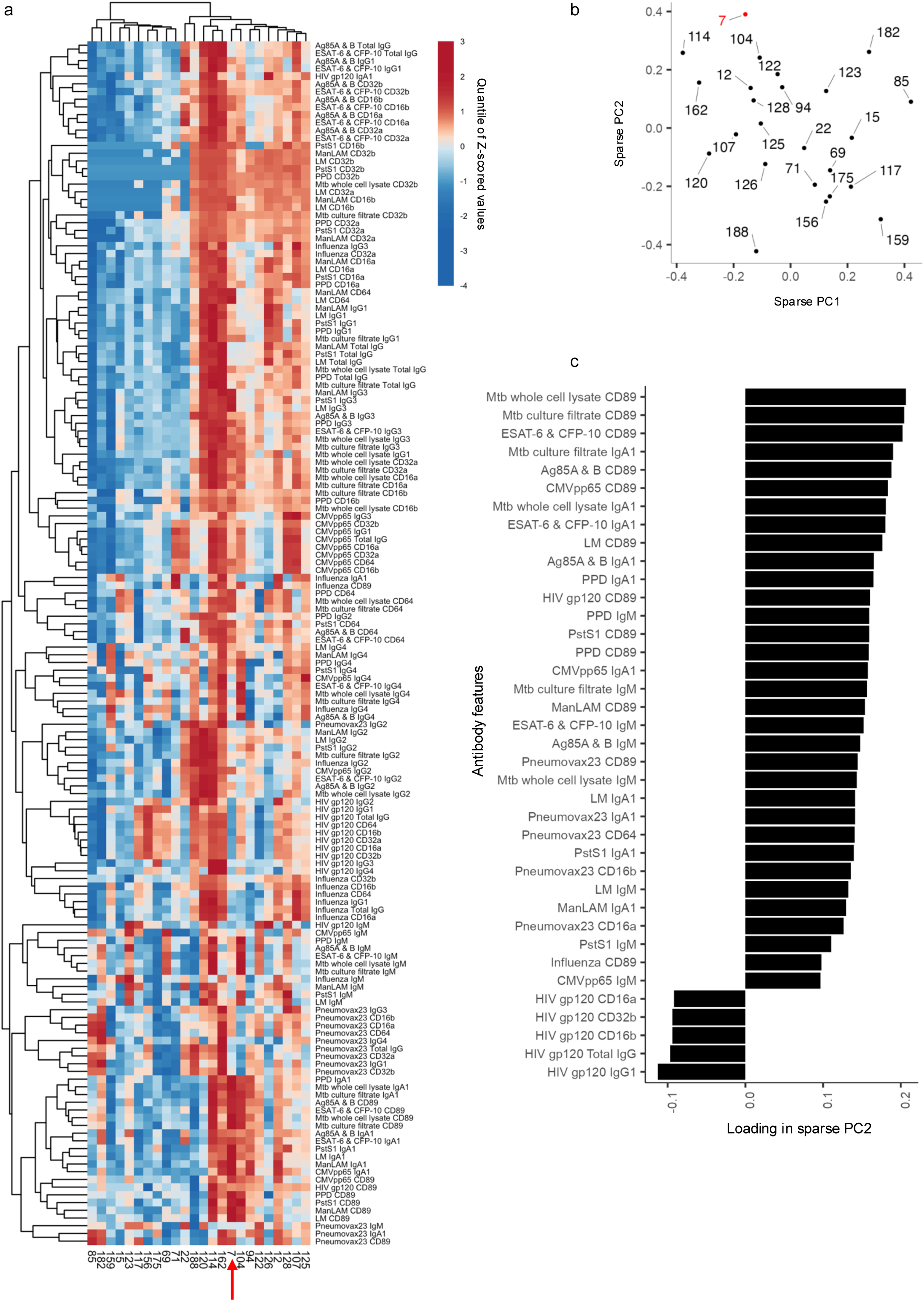
Antigen specific antibody Fc signature in an individual who clinically did not respond to TB treatment. Heatmap shows clustering by individuals (columns) and pre-TB treatment antigen-specific antibody properties and functions (rows) using unsupervised Euclidean distance. The red arrow highlights the individual non-responder to TB treatment (#7) (a). Score plot from sparse principal components analysis of pre-treatment antigen specific antibody properties shows the polyclonal antibody signature of the drug non-responder (red #7) in relationship to other patients (b). Sparse PC2 loadings show what distinguishes the non-responder (red #7) from those responsive to TB treatment (b-c).

**Supplemental Figure 3.**
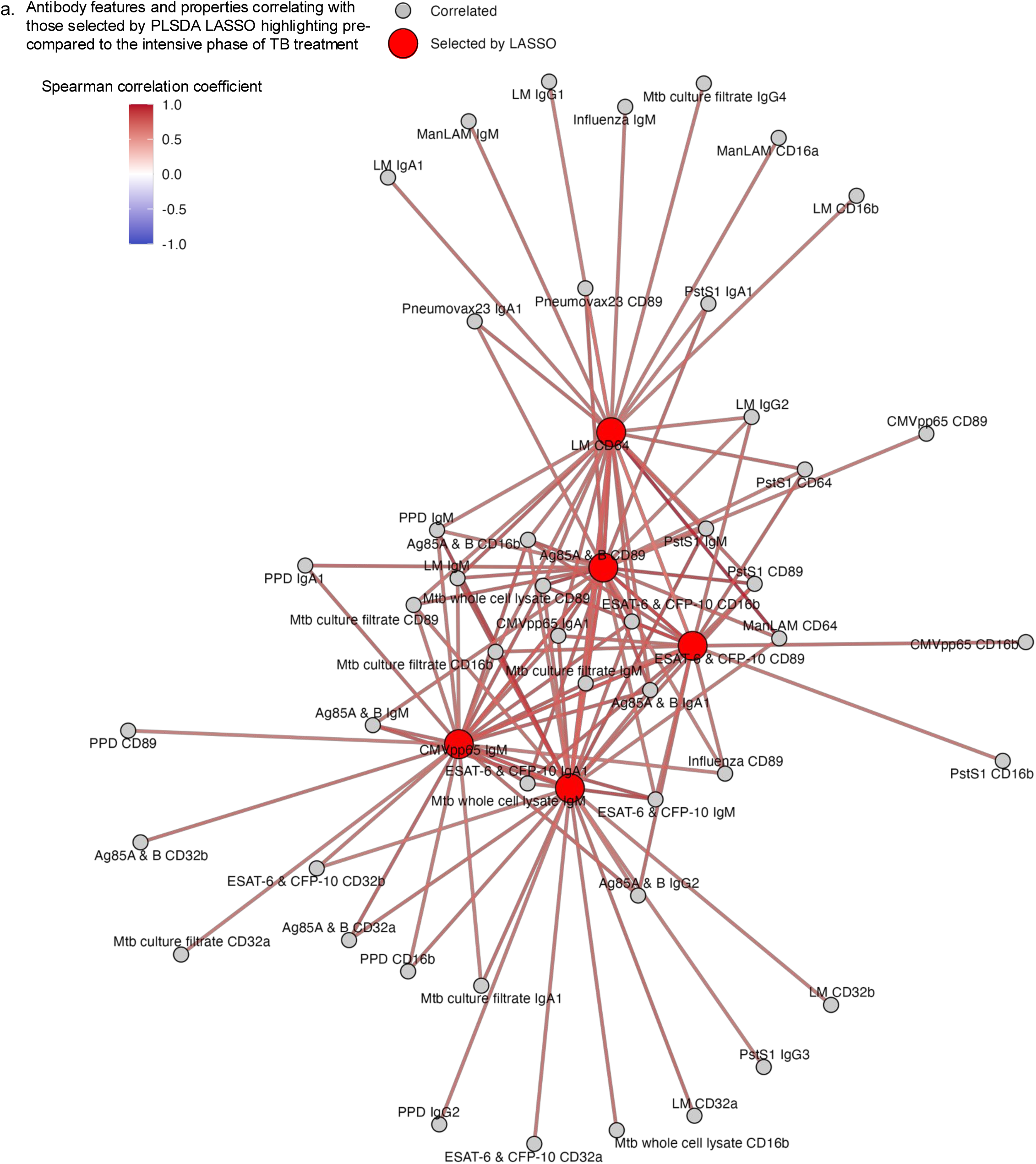
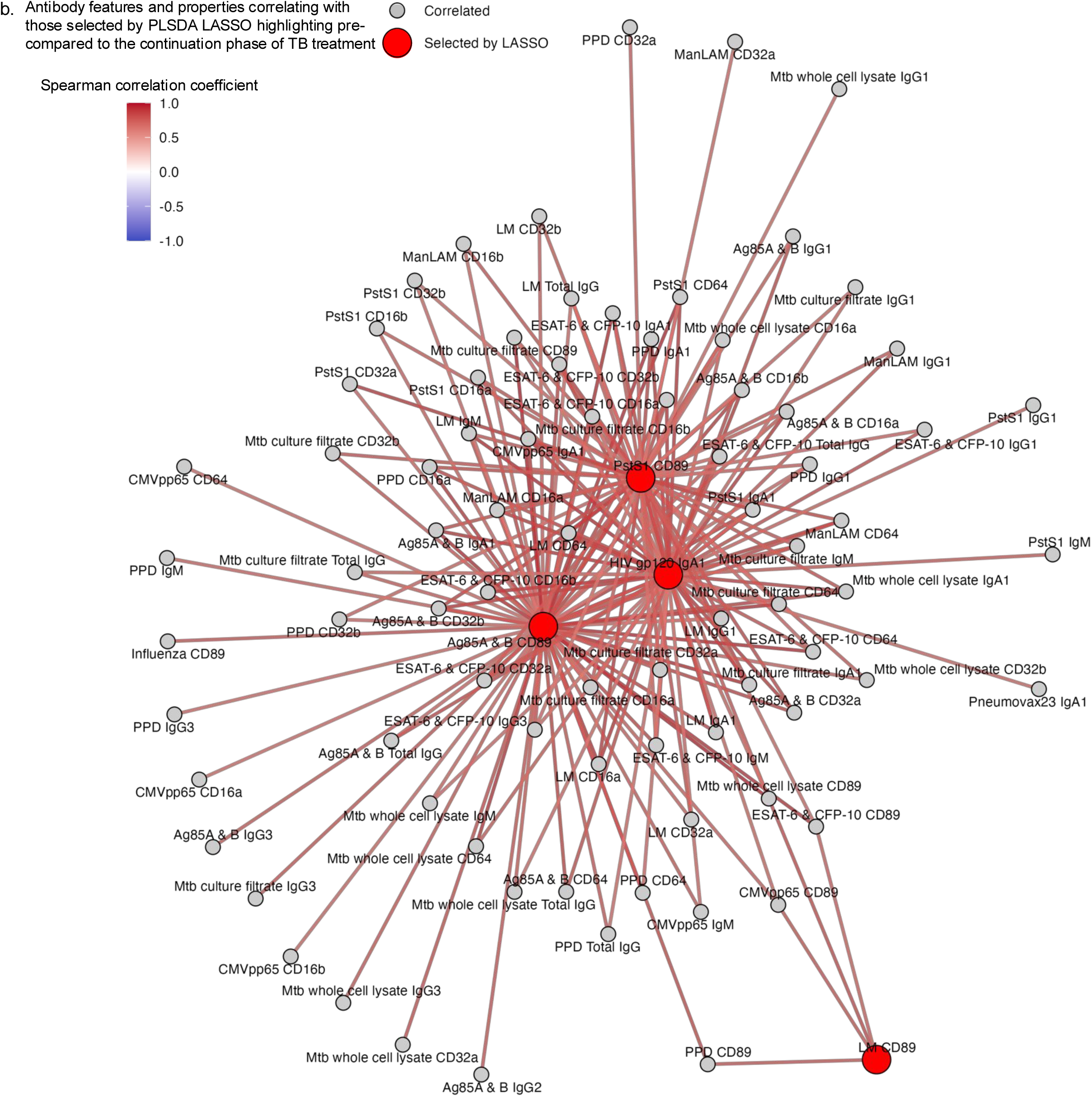
Evolution of antigen specific antibody networks during TB treatment. Network plots depict relationships determined by Spearman correlation between all antigen specific antibody properties and those selected by PLSDA with LASSO in Figure 4 that separate individuals pre-treatment (N=24) from intensive (N=23) (a) and continuation (N=27) (b) phases. Relationships with correlation coefficients≥|0.6| and p<0.05 are shown.

**Supplemental Figure 4.**
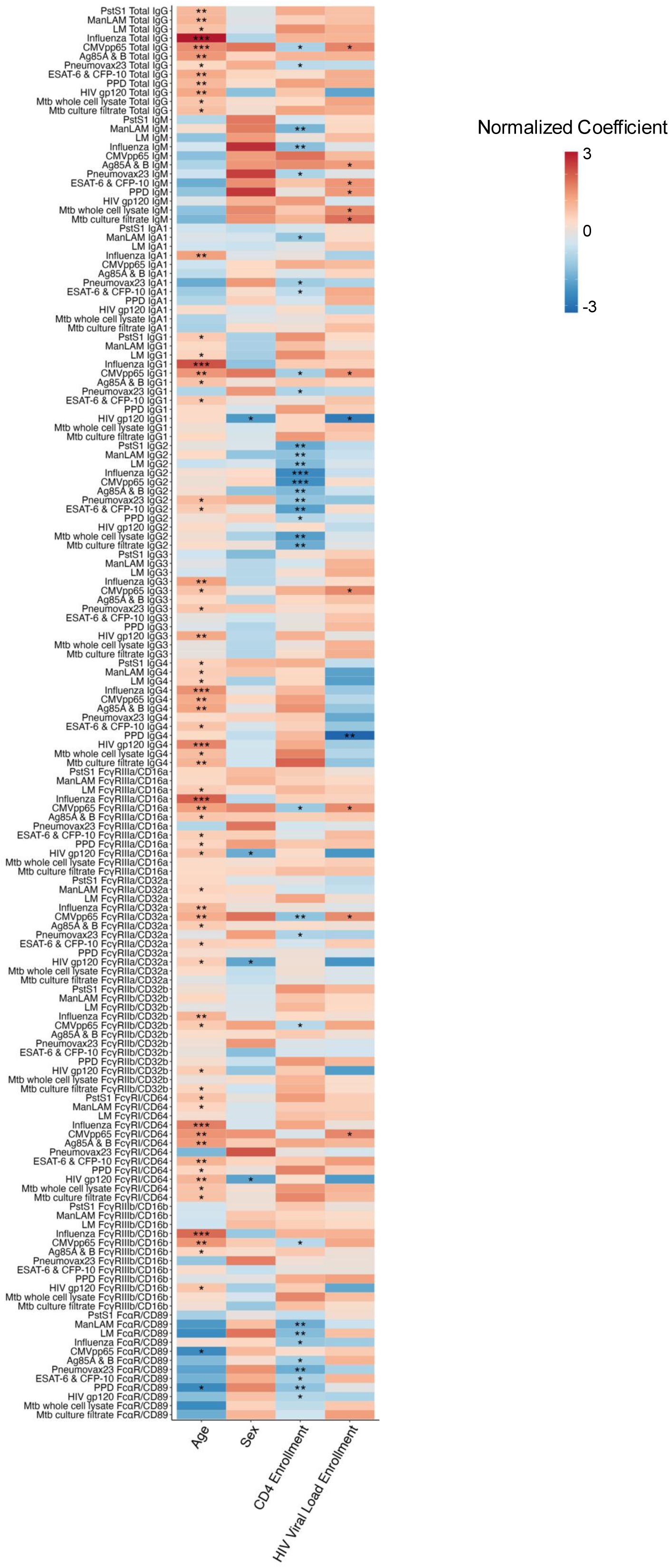
Associations between antigen specific antibody properties and functions with age, sex, CD4, and HIV VL. Heatmap shows the normalized coefficients from multiple linear regressions in Figure 6 assessing the relationship between all pre-TB treatment antigen-specific antibody properties and clinical characteristics for all individuals (N=80). p≤0.05 (*), p≤0.01 (**), and p≤0.001 (***)

## Notes

### Clinical Trial

NCT02063880

### Author Declarations

Ethics committee/IRB of University of Nairobi, Kenyatta National Hospital, University of Washington, Emory University, UT Southwestern Medical Center gave ethical approval for this work.

